# Olfactory Memory Dysfunction in Patients with Traumatic Brain Injury

**DOI:** 10.64898/2026.07.10.26357761

**Authors:** Alefiya Dhilla Albers, Benoît Jobin, Craig A Rovito, Valerie Tseng, Abigail Marshall, Nancy Boudreau, Daniel H Daneshvar, Ross Zafonte, Mark W. Albers

## Abstract

Traumatic brain injury (TBI) severity is typically classified using clinical indices that may have limited prognostic value. Objective measures of olfactory function depend on sensory, limbic, and memory networks and may provide a more specific marker of injury-related neural dysfunction.

Seventy-nine individuals with TBI (48 mild, 31 moderate-to-severe) and 59 healthy controls completed an olfactory battery, including tests of odor percept identification (OPID9, OPID18), odor discrimination (OD10), and odor memory (POEM) ∼4.04 (4.45) years after their most recent TBI. General linear models examined associations between olfactory outcomes and TBI severity, adjusting for age, age², sex, and education. Additional models examined the relationship between loss of consciousness (LOC) and olfactory functioning.

The severity of the most recent TBI was significantly associated with all olfactory outcomes, after adjusting for age, sex, and education. Compared with controls, participants with moderate- to-severe TBI showed lower OPID9, OPID18, POEM, and OD10 performance, while participants with mild TBI showed lower OPID18 and POEM performance. LOC was associated specifically with odor memory in these models, as participants with prolonged (> 30min) LOC or LOC of unknown duration had lower POEM scores than those with no LOC. In TBI-only models, LOC remained associated with POEM after adjustment for TBI severity, whereas TBI severity was not associated with POEM after LOC was included.

Long term olfactory functioning is sensitive to TBI severity, with generalized impairments across odor identification, discrimination, and odor memory. LOC characteristics appear especially relevant to odor memory, suggesting that olfactory memory may capture injury-related features beyond TBI severity classification alone.

## INTRODUCTION

Traumatic brain injury (TBI) is one of the leading causes of death and disability in the United States. The Centers for Disease Control and Prevention estimates there were approximately 223,135 TBI-related hospitalizations in 2019 and 64,362 TBI-related deaths in 2020, with an estimated millions of additional TBIs treated in emergency departments, primary care practices, urgent care clinics, or otherwise untreated.^1^

A clinical assessment of TBI severity currently relies upon instruments like the Glasgow Coma Scale (GCS) and standard forms of brain imaging like a CT scan, which may not be sufficient to capture the complexity and heterogeneity of the condition, as well as objective assessments of brain health and more advanced forms of imaging^2^. Investigating such approaches is critical to advance the development of therapeutics and better diagnostic and prognostic biomarkers of brain injury and recovery over time.

Sensory deficits are common sequelae of TBI, yet TBI’s impact on olfactory function is often overlooked. Olfactory deficits were found to be present in 20.5 million (13.5%) adults 40 years and older in the United States in 2016, with head trauma accounting for 5-17% of these cases.^3,4^ While often regarded as an inconvenience rather than a health concern by patients and physicians alike, olfactory dysfunction reduces quality of life and increases the risk of depression and exposure to environmental hazards.^5^

Patients with TBI commonly develop impaired olfactory function post-injury, with estimates of 5-10% developing anosmia and 30-40% developing hyposmia, depending on the mechanism of head injury.^6^ Olfactory dysfunction can be caused by injuries to any portion of the olfactory pathway, from the superior nasal cavity to the cortical processing centers in the frontal and temporal lobes. Potential neurosensory mechanisms of olfactory dysfunction following head trauma include: 1) damage to the nasal epithelium; 2) shearing of olfactory nerve fibers at the cribriform plate; and 3) direct (e.g., contusion, hemorrhage) or indirect damage to the olfactory bulb, olfactory tract, or regions of cortex involved in odor processing.^3,7^ Emerging evidence suggested that TBI patients with olfactory dysfunction had more damage to brain regions associated with olfaction - including the olfactory bulb, temporal lobe, orbitofrontal cortex, and the left inferior insula - and the level of damage in these regions was correlated with odor discrimination and odor identification ability.^8–10^

Previous research on TBI’s impact on olfactory function has focused on odor identification, odor discrimination, and odor threshold with mixed findings. Green et al. found that odor discrimination was significantly correlated with TBI severity assessed by duration of post-traumatic amnesia (PTA), Glasgow Coma Scale (GCS) score, or abnormalities in neuroimaging, with longer PTA showing the strongest correlation with impaired odor discrimination.^11^ Frasnelli et al. also identified a correlation between PTA duration and odor identification, but failed to find any associations between GCS score and olfactory function.^12^ Similarly, Haxel et al., Neumann et al., and Callahan and Hinkebein did not find significant associations between olfactory dysfunction and TBI severity measured by the GCS.^13–15^ In parallel, no association between either odor identification or odor threshold was found in studies that classified TBI severity based on loss of consciousness (LOC) duration, PTA duration, or neuroimaging results.^15–17^ Given the mixed evidence supporting a relationship between olfactory dysfunction and TBI severity based on single measures, a multidimensional clinical classification of TBI severity, incorporating LOC duration, PTA duration, and GCS score, may better account for variability in olfactory dysfunction across the TBI severity spectrum.

Impairment in odor memory has been documented in preclinical and early stages of neurodegenerative diseases, such as Alzheimer’s disease (AD) and Parkinson’s disease.^18–21^ Repeated TBI, most common in contact sports athletes, is a major risk factor for the development of these diseases and other neurodegenerative conditions.^22^ Despite the causal relationship between TBI and neurodegeneration and the observed impairments in odor memory in neurodegenerative disease, deficits in odor memory remain to be explored in the context of TBI. The strong anatomical connections between the primary olfactory cortex and hippocampus may explain the intimate relationship between olfaction and memory, best embodied by odor-evoked memory.^23,24^

The objective of this study was to assess odor percept identification (OPID), odor discrimination (OD), and percepts of odor episodic memory (POEM), measured by a novel olfactory battery, across TBI severity levels classified by a clinical metric of multiple TBI-related variables. An odor percept is the mental impression of an experienced odor isolated from other contextual information. During the test, an odor is presented to a subject before the response choices are given. This procedure provides no visual or semantic information to allow the subject to contextualize the odor. Subjects must retain the percept of an odor in working memory to complete each test. To our knowledge, this is the first olfactory test incorporating working memory into an odor identification task and episodic memory of odor percepts in those with TBI.

Episodic odor memory has been experimentally investigated in behavioral and functional neuroimaging studies reporting the crucial role of the piriform cortex and hippocampus.^24^ We previously incorporated episodic odor memory into a similar psychophysical test of olfactory function, and illustrated that a specific deficit in odor memory correlated with AD biomarkers in a preclinical population.^20^

## MATERIALS AND METHODS

### Participants

This study focused on subjects who received outpatient or inpatient care for TBI at Massachusetts General Hospital (MGH) and/or Spaulding Rehabilitation Hospital (SRH) from 2000 to October 2019.

Potential participants were contacted by their physician with a recruitment letter and study flyer, and followed up by phone for scheduling if interested. Healthy age-matched control participants were recruited from the community. Prescreening for eligibility criteria was conducted by reviewing electronic medical records (for inpatients and outpatients) and during the follow-up phone call. During the study visit, informed consent was obtained in writing by an Institutional Review Board (IRB)/CITI-certified and HIPAA-trained member of the study staff. The study protocol was approved by the IRB of Mass General Brigham.

### Eligibility criteria

Female and male patients with a history of diagnosis of TBI were eligible to participate in the study. TBI severity was determined using all available information about the most recent TBI from the electronic medical record, the Demographics/Medical History form, and, for participants enrolled through SRH, data obtained as part of the TBI Model Systems National Database. Moderate-to-severe TBI was classified by the presence of at least one of the following criteria: PTA lasting more than 24 hours, LOC exceeding 30 minutes, GCS less than 13, or structural neuroimaging abnormalities. All other TBIs were classified as mild. When a severity indicator was missing, it was not used for classification; participants were not excluded solely because one or more severity indicators were unavailable, provided that sufficient available clinical information supported classification. In the present sample, mild TBI participants had either no LOC (n = 30), brief LOC <30 minutes (n = 13), or LOC of unknown duration (n = 5). Among mild TBI participants with non-missing PTA data, 5 reported PTA and 23 did not. Among mild TBI participants with non-missing GCS data, all had a score of 15. Among mild TBI participants with non-missing neuroimaging data, all had normal findings.

Patients with TBI and controls included in the study were 18 years or older. Further exclusion criteria included: 1) traumatic or congenital anosmia; 2) current sinus or upper respiratory infection; 3) nasal polyps; 4) trouble breathing due to deviated septum; 5) current or recent (past 6 months) alcohol or substance dependence including smoking; 6) severe cognitive dysfunction; 7) primary pulmonary disease, such as asthma or severe emphysema, not under good medical control; 8) pneumocephalus; 9) basilar skull fracture; 10) CSF leak; 11) facial fractures to include Lefort I, II or III; 12) pregnancy; and 13) preferred language listed other than English.

### Study measures

Demographics and Medical History Form

Demographic data, including age, sex, race/ethnicity, years of education, and exposure to contact sports in a competitive setting, as well as medical factors that may confound olfactory function (nasal, sinus, and pulmonary diseases, deviated septum, history of radiation or chemotherapy), were collected by study staff in writing during the 10-minute break between the first (OPID9) and second (POEM/OPID18) olfactory tests. In addition, self-reported head injury-related information was collected, including the total number of brain injuries, time since last injury, mechanism of each head injury, duration of LOC, duration of PTA, neurosurgical history, and neuroimaging results. When available, this self-report information was validated against the contents of the electronic health record. Subjects were also asked whether they suffered from smell problems, and if so, when these problems emerged.

### Olfactory assessment

The olfactory battery ^20,25^ uses customizable, portable odor-delivery devices called “Whispis” (Scentovation, Portland, ME). Odor stimuli dispensed by the Whispis are part of the Living Library^™^ of odors (International Flavors and Fragrances, New York), and are designed to evoke a naturalistic odor percept. Olfactory testing took approximately 45 minutes to complete, and all data were recorded using the secure RedCap platform. The testing protocol follows and is illustrated in Figure 1,

1. Odor identification (OPID9: 9 trials): After sampling each odor, the subject was asked whether the odor was generally familiar to them. Then subjects were presented with 4 odor names and asked to identify the name that best matched the odor sampled. The OPID9 score was calculated as a proportion of odors named correctly /9. A score of 0.25 is indicative of scoring at the level of chance.
2. Odor Memory & Identification (POEM & OPID18: 18 trials): After sampling each odor, the subject was asked whether the odor was presented previously in the first OPID9 test. Then subjects were presented with 4 odor names and asked to identify the name that best matched the odor sampled. The OPID18 score was calculated as a proportion of odor named correctly /18. A score of 0.25 is indicative of scoring at the level of chance. The POEM index score was calculated as follows: (# correct recognition - # incorrect recognition)/total. Scores range from -1 to 1, with a score of 0 indicative of scoring at the level of chance.
3. Odor Discrimination (OD10: 10 trials): Subjects serially sampled pairs of odors in each trial. Subject was asked whether the pair of odors smelled the same or different. The OD score was calculated as the proportion of trials correctly discriminated as same or different /10. A 0.5 score is indicative of scoring at the level of chance.

**Figure 1:**
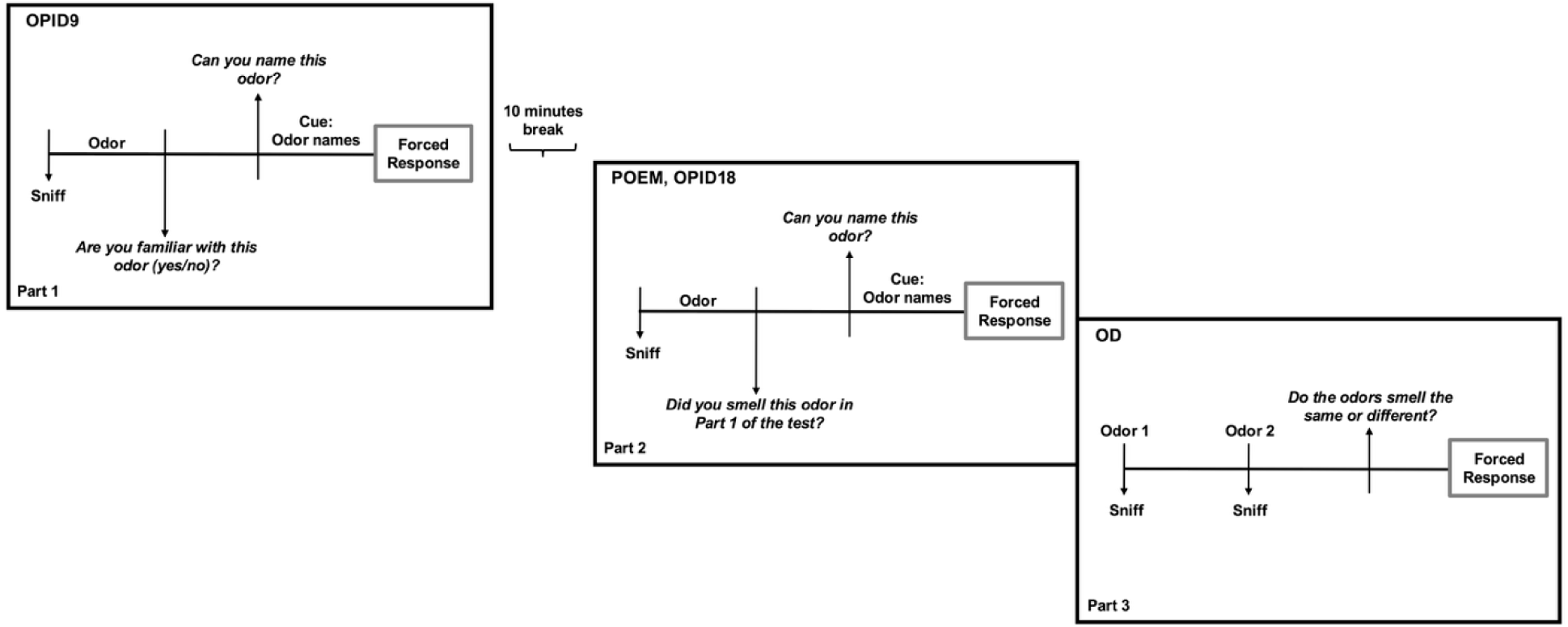
Testing protocol for the olfactory battery. The olfactory assessment was administered in three parts. Part 1: Odor Percept Identification (OPID9). Participants sampled odors and were first asked whether the odor was familiar (yes/no). They were then presented with four odor name options and asked to identify the one that best matched the odor (forced-choice). Part 2: Odor Memory and Odor Percept Identification (POEM & OPID18). Participants sampled additional odors and were asked whether each odor had been presented previously during Part 1 (recognition memory). They then completed a four-alternative forced-choice odor identification task (OPID18). Part 3: Odor Discrimination (OD). Participants serially sampled pairs of odors and indicated whether the two odors smelled the same or different (forced-choice).

### Statistical analysis

Descriptive characteristics were summarized by TBI severity group: control, mild TBI, and moderate-to-severe TBI. Continuous variables were reported as means and standard deviations, and categorical variables were reported as frequencies and percentages. Group differences were examined using Kruskal–Wallis tests for continuous variables and Fisher’s exact tests for categorical variables. Injury-related variables were compared between the mild TBI and moderate-to-severe TBI groups only.

Primary analyses examined the association between TBI severity and olfactory performance. Separate general linear models were fitted for each olfactory outcome: OPID9, OPID18, POEM, and OD10. TBI severity was entered as a three-level categorical predictor: control, mild TBI, and moderate-to-severe TBI. All primary models were adjusted for age, age², sex, and years of education. Type III F tests were used to evaluate the omnibus effect of TBI severity. When the omnibus effect was significant, estimated marginal means and 95% confidence intervals were computed, and Tukey-adjusted pairwise comparisons were used to compare TBI severity groups.

Secondary analyses were restricted to participants with TBI and examined the association between LOC and olfactory performance. LOC was modeled as a four-level categorical variable: no LOC, brief LOC (<30 minutes), prolonged LOC (≥30 minutes), and LOC of unknown duration. Separate general linear models were fitted for OPID9, OPID18, POEM, and OD10. These models were also adjusted for age, age², sex, years of education, number of injuries, and time since last injury. Type III F tests were used to evaluate the omnibus effect of LOC group. When LOC group was significant, estimated marginal means and 95% confidence intervals were computed, and Tukey-adjusted pairwise comparisons were used.

We performed an additional TBI-only analysis to examine whether LOC accounted for the association between TBI severity and odor memory. First, the association between TBI severity and LOC group was tested using Fisher’s exact test. Then, POEM was modeled as a function of TBI severity, adjusted for age, age², sex, education, and number of injuries. LOC group was then added to this model to determine whether TBI severity remained associated with POEM after accounting for LOC, and whether LOC remained associated with POEM after adjustment for TBI severity.

As a secondary analysis among participants with TBI, time since last TBI and number of injuries were examined in separate linear models for each olfactory outcome, adjusted for age, age², sex, education, and number of injuries or time since last TBI, respectively.

Covariate effects from the primary models were also examined, focusing on age, age², and education. Model diagnostics were inspected to evaluate residual distributions and influential observations. Analyses were conducted in R using general linear models, type III tests, estimated marginal means, and Tukey-adjusted post hoc comparisons.

## RESULTS

### Demographics

A total of 79 patients with TBI and 59 controls completed the olfactory battery. Of the 79 patients with TBI, 48 were classified as mild TBI and 31 as moderate-to-severe TBI. Demographic characteristics, including age, sex, race, and education, did not significantly differ across the three groups in unadjusted analyses (Table 1). Among participants with TBI, the mild and moderate-to-severe TBI groups did not significantly differ in the number of TBIs sustained or in time since the most recent injury. As expected, injury-severity indicators differed between TBI groups: compared with the moderate-to-severe TBI group, the mild TBI group had higher available GCS scores and lower proportions of LOC, PTA, and abnormal neuroimaging findings. Because some clinical severity indicators were missing for a subset of participants, Table 1 reports row-specific available data. Missingness in PTA, GCS, or neuroimaging status did not exclude participants from the primary olfactory analyses, which were based on the final clinical TBI severity classification.

**Table 1.**
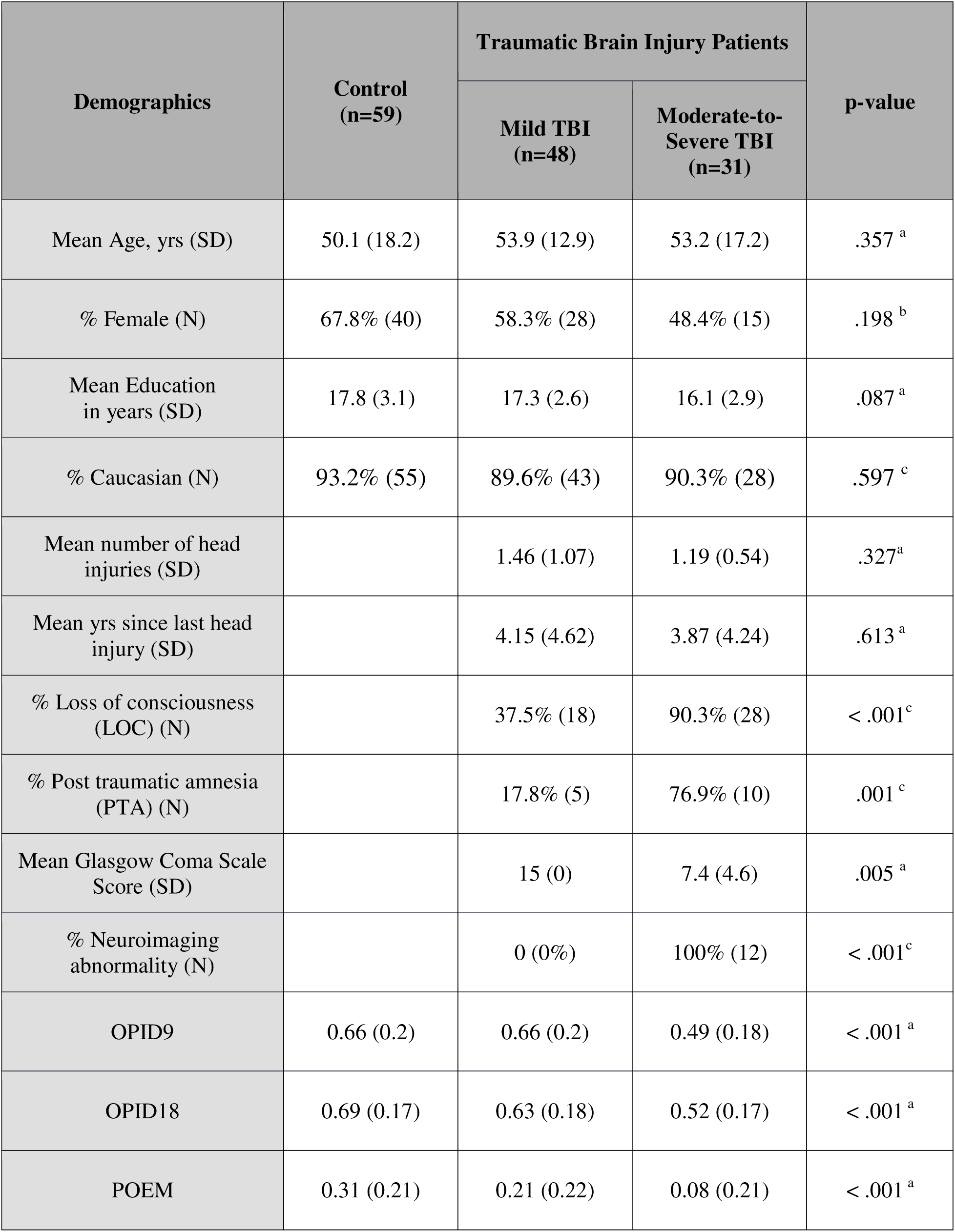
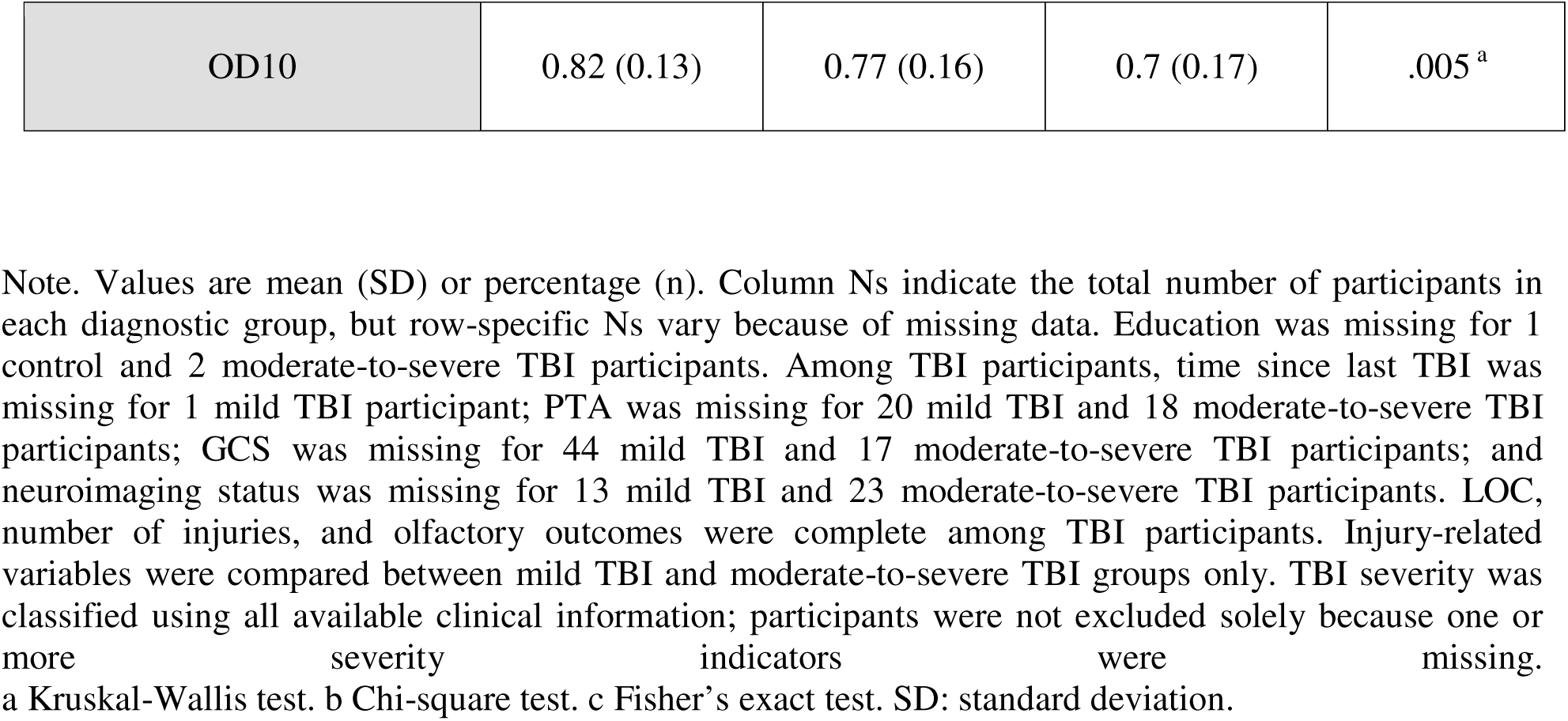
Summary of demographics and head injury-related variables in the cohort.

### TBI severity

TBI severity was significantly associated with olfactory scores across all outcomes after adjustment for age, age², sex, and education (Table 2; Figure 2). For odor identification, TBI severity was significantly associated with both OPID9, *F*(2, 128) = 6.58, *p* = .002, ηp² = .093, and OPID18, *F*(2, 128) = 8.71, *p* < .001, ηp² = .120. Tukey-adjusted post hoc comparisons showed that participants with moderate-to-severe TBI performed worse than controls on OPID9, Δ = 0.155, *p* = .002, and OPID18, Δ = 0.159, *p* < .001. Participants with mild TBI also performed worse than controls on OPID18, Δ = 0.082, *p* = .042, whereas the mild TBI and moderate-to-severe TBI groups did not significantly differ from each other.

**Table 2.**
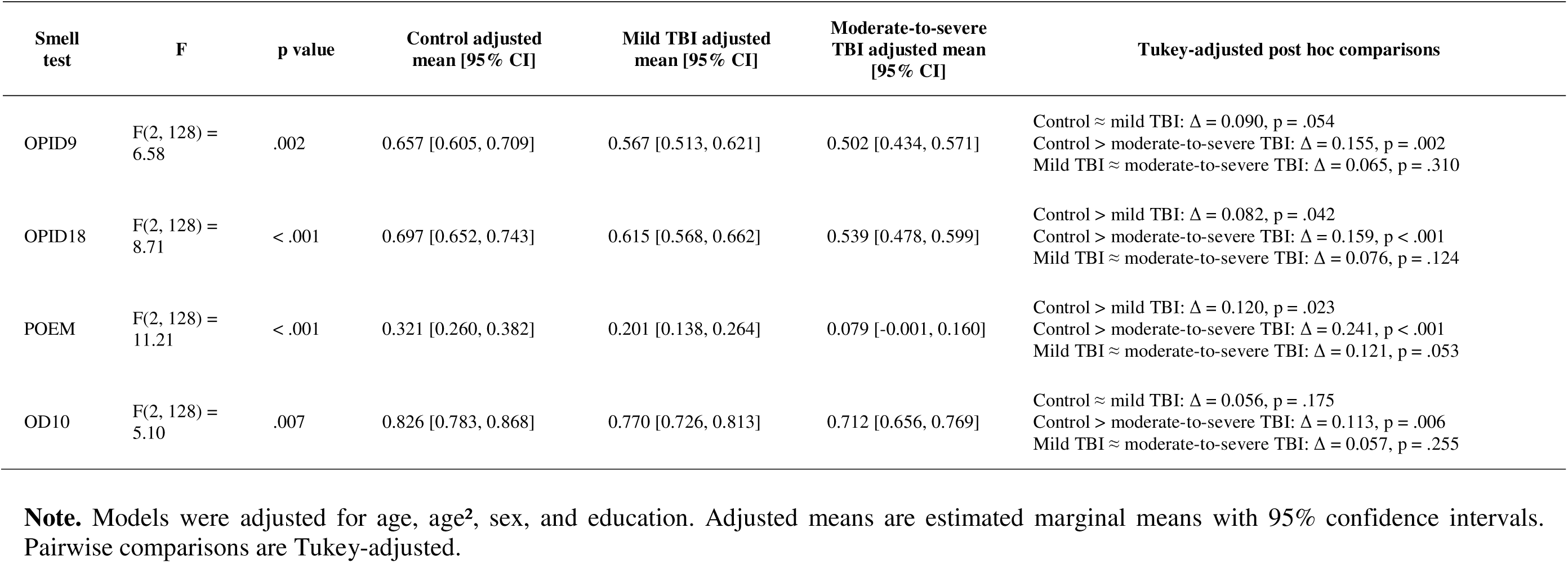
TBI severity and olfactory outcomes.

**Figure 2.**
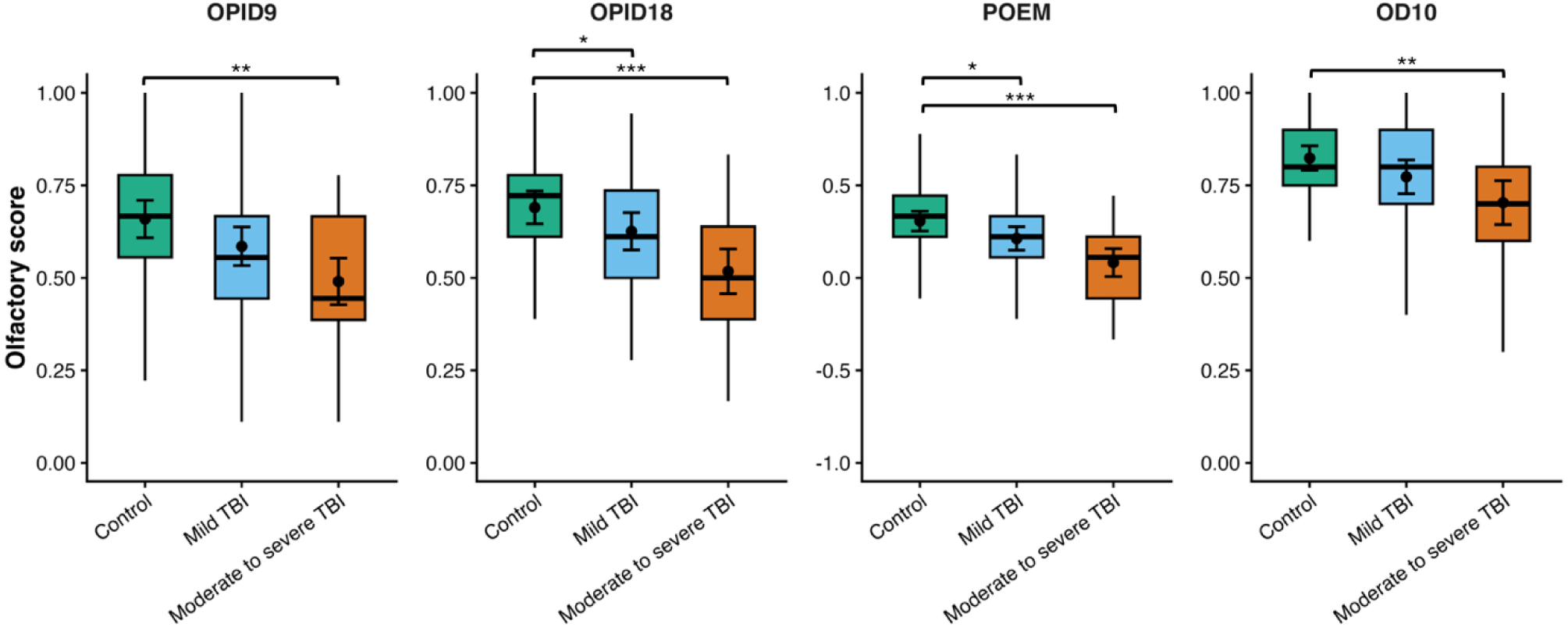
Olfactory performance by TBI severity. Boxplots show the median and interquartile range for each TBI severity group. Black points represent group means, and error bars represent 95% confidence intervals. * *p* <.05 ** *p* <.01 *** *p* <.001

TBI severity was also significantly associated with odor memory (POEM) *F*(2, 128) = 11.21, *p* < .001, ηp² = .149. Post hoc comparisons indicated that both the mild TBI group, Δ = 0.120, *p* = .023, and the moderate-to-severe TBI group, Δ = 0.241, *p* < .001, had lower adjusted POEM scores than controls. Finally, TBI severity was significantly associated with odor discrimination, OD10, *F*(2, 128) = 5.10, *p* = .007, ηp² = .074, with lower performance in the moderate-to-severe TBI group compared with controls, Δ = 0.113, *p* = .006.

### Loss of consciousness

Among participants with TBI, LOC group (no LOC, brief LOC <30 minutes, prolonged LOC ≥30 minutes, and LOC of unknown duration) was significantly associated with odor memory performance (POEM, *F*(3, 66) = 5.58, *p* = .002, ηp² = .202), but not with odor identification or odor discrimination (Figure 3). Adjusted POEM scores were highest among participants with no LOC (M = 0.259, 95% CI [0.182, 0.336]), followed by those with brief LOC <30 minutes (M = 0.182, 95% CI [0.088, 0.277]), LOC of unknown duration (M = 0.038, 95% CI [−0.096, 0.173]), and prolonged LOC ≥30 minutes (M = −0.006, 95% CI [−0.127, 0.115]). Tukey-adjusted post hoc comparisons indicated that participants with no LOC had significantly higher POEM scores than those with prolonged LOC, Δ = 0.265, *p* = .003, and those with LOC of unknown duration, Δ = 0.220, *p* = .031. The difference between brief LOC and prolonged LOC was in the expected direction but did not reach statistical significance, Δ = 0.188, *p* = .081. No other pairwise comparisons were statistically significant.

**Figure 3.**
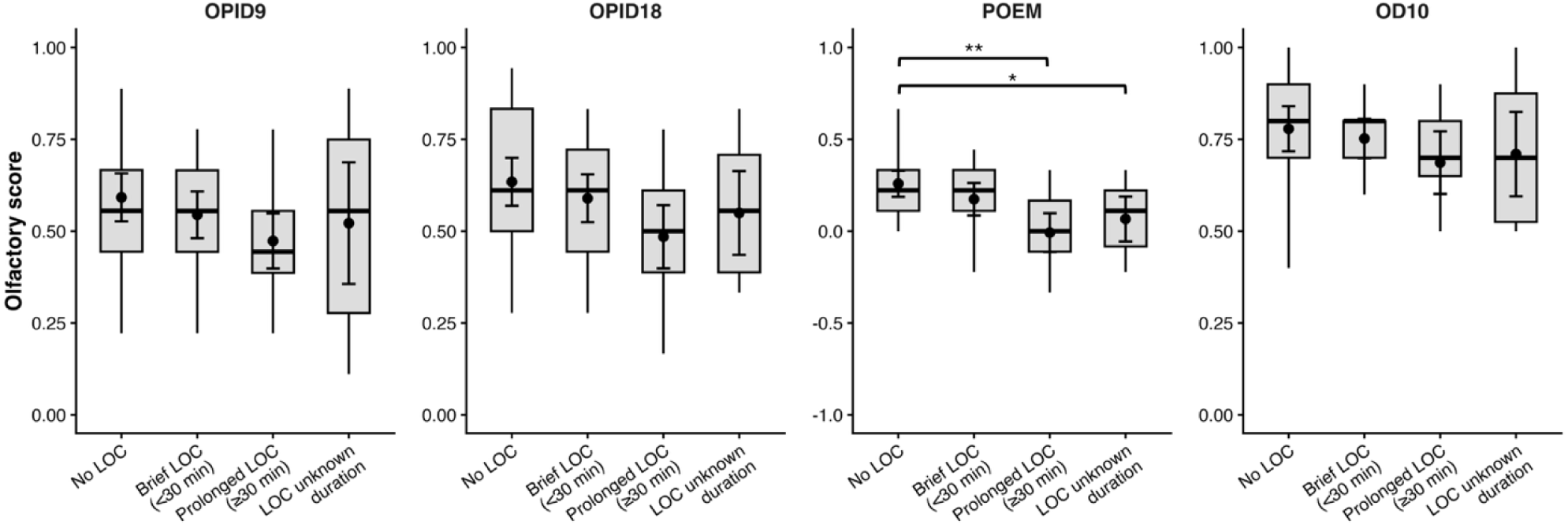
Olfactory performance by loss-of-consciousness duration among patients with TBI. Boxplot show the median and interquartile range for each LOC duration group. Black points and error bar represent raw group means and 95% confidence intervals. * *p* <.05 ** *p* <.01

**Figure 4.**
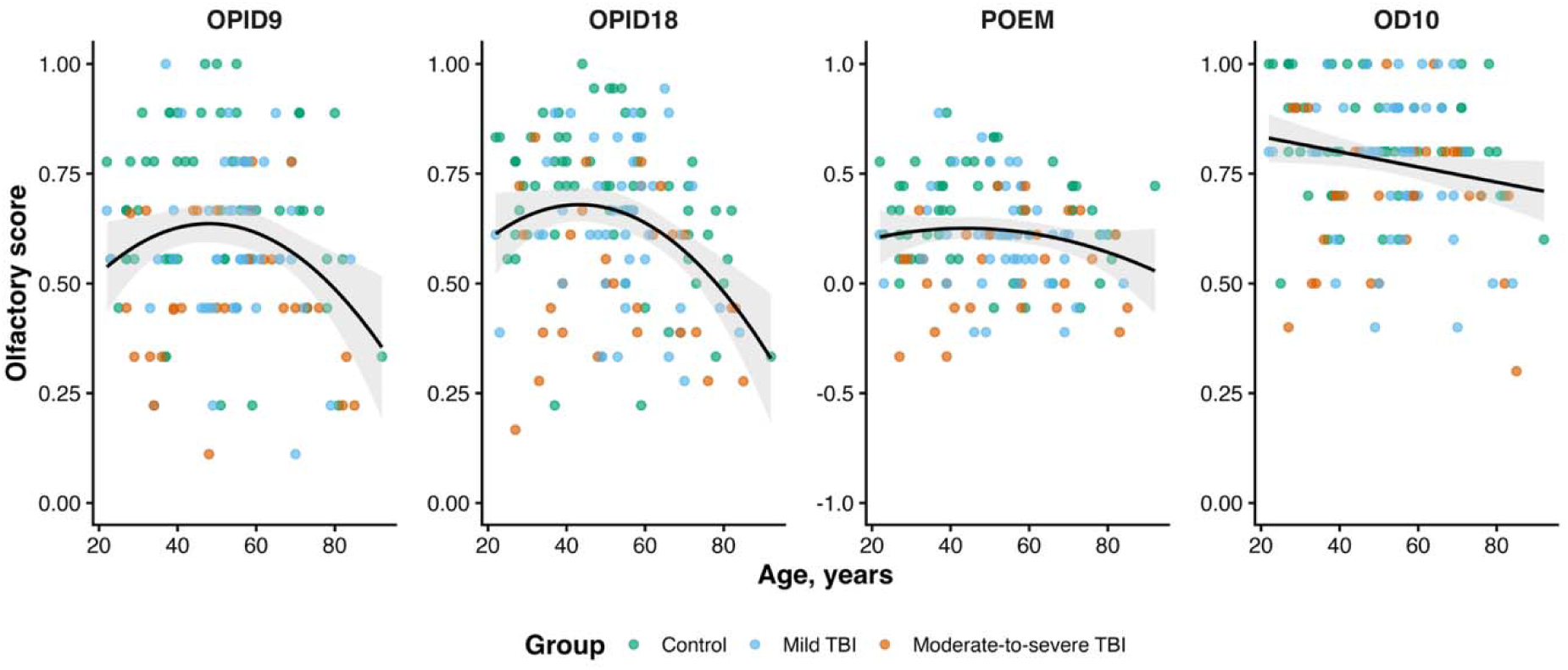
Relationship between age and olfactory performance. Points represent raw individual scores. Black lines show fitted age effects with 95% confidence bands. OPID9, OPID18, and POEM were fit with quadratic age trends, whereas OD10 was fit with a linear age trend, consistent with the retained model terms. Only OPID9 and OPID18 associations with age were statistically significant.

LOC was not significantly associated with OPID9, *F*(3, 66) = 0.59, *p* = .622, ηp² = .026, OPID18, *F*(3, 66) = 1.80, *p* = .157, ηp² = .076, or OD10 scores, *F*(3, 66) = 0.48, *p* = .698, ηp² = .021. Thus, within the TBI group, LOC severity appeared most strongly related to odor memory rather than odor identification or odor discrimination.

Because both LOC and TBI severity showed associations with POEM, we examined whether each remained associated with odor memory when included in the same model among participants with TBI. TBI severity and LOC group were strongly associated with each other, Fisher’s exact test with simulated p-value, *p* < .001. In a model adjusted for age, age², sex, education, number of injuries, and time since last injury, TBI severity showed a trend-level association with POEM, *F*(1, 68) = 3.23, *p* = .077, ηp² = .045. When LOC group was added to the model, TBI severity was no longer associated with POEM, *F*(1, 65) = 0.60, *p* = .442, ηp² = .009, whereas LOC group remained significantly associated with POEM, *F*(3, 65) = 4.50, *p* = .006, ηp² = .172. These findings suggest that LOC characteristics may be more closely related to odor memory performance than overall TBI severity among participants with TBI.

### Time since last TBI

Mean time since last TBI was similar across TBI severity groups: 4.15 ± 4.62 years in the mild TBI group (range: 0.18–19.24 years) and 3.87 ± 4.24 years in the moderate-to-severe TBI group (range: 0.02–14.08 years). Mean time since last TBI was similar across TBI severity groups: 4.15 ± 4.62 years in the mild TBI group and 3.87 ± 4.24 years in the moderate-to-severe TBI group. Time since last TBI was not significantly associated with OPID9, *F*(1, 69) = 1.90, *p* = .173, ηp² = .027, OPID18, *F*(1, 69) = 1.07, *p* = .305, ηp² = .015, POEM, *F*(1, 69) = 0.59, *p* = .446, ηp² = .008, or OD10, *F*(1, 69) = 2.27, *p* = .137, ηp² = .032.

### Number of injuries

Number of injuries was not significantly associated with any olfactory outcome among participants with TBI after adjustment for age, age², sex, education, and time since last injury. Specifically, number of injuries was not associated with OPID9, *F*(1, 69) = 0.21, *p* = .645, ηp² = .003, OPID18, *F*(1, 69) = 0.00, *p* = .956, ηp² < .001, POEM, *F*(1, 69) = 2.53, *p* = .117, ηp² = .035, or OD10, *F*(1, 69) = 1.37, *p* = .246, ηp² = .019. These findings suggest that, within the TBI group, the number of reported injuries was not a robust predictor of olfactory performance.

### Covariate effects

Covariate effects were examined in the fully adjusted TBI-only models that included TBI severity, number of injuries, time since last injury, age, age², sex, and education. For OPID9, there was a significant quadratic effect of age, *F*(1, 68) = 5.30, *p* = .024, ηp² = .072, whereas the linear age term and education were not significant. For OPID18, the quadratic age term was significant, *F*(1, 68) = 5.73, *p* = .019, ηp² = .078, whereas the linear age term was not significant. Education showed a trend-level association with OPID18, *F*(1, 68) = 3.54, *p* = .064, ηp² = .049.

For POEM, the quadratic age effect was significant, *F*(1, 68) = 4.56, *p* = .036, ηp² = .063, whereas the linear age term and education were not significant. For OD10, the quadratic age effect was also significant, *F*(1, 68) = 5.61, *p* = .021, ηp² = .076, whereas the linear age term wa not significant. Education showed a trend-level association with OD10, *F*(1, 68) = 3.42, *p* = .069, ηp² = .048.

## Discussion

To our knowledge, this study represents the first analysis of odor memory in patients with a history of TBI. We found that TBI severity was associated with broad olfactory impairment, including lower odor identification, odor discrimination, and odor memory performance, with the largest deficits generally observed in participants with moderate-to-severe TBI relative to healthy controls. In contrast, among participants with TBI, LOC characteristics were specifically associated with odor memory, whereas time since last TBI was not significantly related to olfactory performance. These findings suggest that a broad clinical categorization of increased TBI severity associates with decreased olfactory function across a variety of domains (identification, memory, discrimination), while a more specific clinical measure like LOC associates with a more specific and central olfactory measure like odor memory.

While the categorization of brain injuries may be supplemented by olfactory measures in the moderate and severe realm, olfactory outcomes could provide additional value as a diagnostic tool for mild TBI (mTBI). The characterization and definition of mTBI have largely relied on subjective symptom scores and observation of difficult to ascertain concussions signs, resulting in substantial debate over decades.^26–29^ Some concussion signs and symptoms can be subjective, vague, or be related to other causes, complicating diagnostic decision making, especially in those with multiple concussions or with persistent post-concussion symptoms. An objective psychophysics olfactory test, which includes odor memory, could also assist clinicians in these more symptomatically nuanced cases of mTBI. As an illustration of its clinical relevance and potential as a prognostic indicator, prior work showed that olfactory impairment following an mTBI was associated with persistence of symptoms of depression and/or anxiety six months later. ^30^

In the broader mTBI literature, LOC has been associated with poorer prospective memory among patients with mTBI^31^, with lower memory performance one month after injury accompanied by reduced bilateral hippocampal volume six months later^31^, suggesting that disruption of consciousness may index injury mechanisms relevant to later memory functioning. This state of paralytic coma is associated with a longer recovery time and more severe brain injury, specifically subcortical contusions and deeper lesions by neuroimaging.^32^ Because odor memory relies on olfactory–limbic circuitry, including medial temporal structures such as the hippocampus and entorhinal cortex, LOC may be particularly relevant to POEM performance by reflecting transient disruption or injury to such memory-supporting networks. A meta-analysis of studies examining the clinical impacts of LOC in mTBI indicates an increased risk of negative outcomes like post-concussive syndrome, post-traumatic stress disorder, depression, and a decreased quality of life.^33^ These results are intriguing given the specific association we found between LOC and odor memory, and suggest that deficits in odor memory after TBI may serve as a useful prognostic biomarker for LOC-associated clinical outcomes. Our results position odor memory as a potential post hoc indicator of LOC, in addition to clinical reporting of LOC symptoms, which could be underreported depending on the severity/duration of LOC.

Age showed an inverted–U–shaped quadratic association with both odor percept identification and odor memory, whereas odor discrimination demonstrated a negative linear relationship with age. Specifically, the predictive model indicated that younger and older individuals performed worse on odor identification and memory tasks compared with middle-aged participants, consistent with an inverted-U–shaped lifespan trajectory commonly observed for tasks relying on semantic knowledge and controlled semantic retrieval.^34,35^ This similar pattern observed is not surprising, as odor identification involves declarative memory processes and is associated with declarative memory performance involving semantic and episodic processes.^36,37^ Although the mechanisms underlying this pattern are not fully understood, one plausible explanation relates to odor familiarity and cumulative semantic experience of odors. Semantic knowledge typically increases from early adulthood into midlife before stabilizing or declining, reflecting lifelong odor exposure and percept learning.^38^

Importantly, our findings suggest that TBI severity is associated with a generalized olfactory impairment, with reductions in odor identification, odor discrimination, and odor memory compared to controls. This pattern of olfactory impairment observed years after the most recent TBI differs from the profile typically described in prodromal Alzheimer’s disease (AD). Whereas prodromal AD is characterized by early and specific odor identification and odor memory impairment linked to medial temporal lobe regions^20,21,39,40^, TBI-related olfactory dysfunction may arise from more heterogeneous mechanisms, including shearing of olfactory nerve fibers at the cribriform plate, damage to the olfactory epithelium or olfactory bulb, and diffuse injury to primary and secondary olfactory brain regions.^3^ This broader pattern may resemble, in some respects, anosmia observed in Parkinson’s disease, where early pathology involving the olfactory bulb and anterior olfactory structures contributes to generalized smell deficits.^41,42^

The limitations of this study include lack of diversity. Population-level data indicate that TBI-related emergency department visits vary by age, sex, and race/ethnicity, with higher rates among males, Black individuals, and individuals at the extremes of age. This differs from the present sample, which was predominantly Caucasian, female, and around 50 years of age, and therefore may not represent the broader population of patients with moderate-to-severe TBI. To be broadly applied to the TBI population, these findings should be replicated in a more diverse and representative sample. Additional limitations include the modest sample size and heterogeneity of TBI. Although 79 participants with TBI enabled the study to examine group-level associations, a larger sample would better capture variability in injury mechanisms, lesion locations, clinical interventions such as craniotomy or craniectomy, and cumulative injury exposure. In addition, some clinical injury indicators, particularly PTA, GCS, and neuroimaging status, were not available for all participants, reflecting the use of real-world clinical and historical data across recruitment sources where injury documentation can vary. Importantly, the main variables used in the present analyses were highly complete: LOC, number of injuries, and all olfactory outcomes were available for all participants with TBI, and time since last TBI was missing for only one participant. Thus, missingness primarily affected the descriptive characterization of some injury-severity indicators rather than the main olfactory analyses.

To conclude, this study showed that TBI severity was associated with a generalized olfactory impairment pattern, with poorer odor identification, odor discrimination, and odor memory.

Among participants with TBI, LOC was most specifically related to odor memory performance even after adjusting for TBI severity. In contrast, time since last TBI was not significantly associated with olfactory outcomes, suggesting that these olfactory differences may reflect long-term injury-related features rather than simply time elapsed since injury. Future prospective studies with standardized acute injury documentation and neuroimaging would allow more precise characterization of the relationships among olfactory function, injury severity, injury mechanisms, and cognitive outcomes.

## TRANSPARENCY, RIGOR, AND REPRODUCIBILITY STATEMENT

This clinical study was not preregistered due to its observational and exploratory nature. The analysis plan was not formally preregistered but the lead author (A.D.A.) certifies that the analysis plan was pre-specified. A sample size of 150 subjects was planned based on availability: 50 per group (mild TBI, moderate-to-severe TBI, healthy controls). The actual sample size was 79 participants with traumatic brain injury (48 mild, 31 moderate-to-severe) and 59 healthy controls who completed a battery of olfactory tests approximately 4.04 ± 4.45 years after their most recent TBI. The observed effect sizes of TBI severity on olfactory performance were medium to large (ηp² = .074–.149). 138 participants were screened from Massachusetts General Hospital, Spaulding Rehabilitation Hospital, and the community and all participated in and completed study procedures. Participants were not told the results of their olfactory assessments.

Data collection and analyses were performed by team members blinded to relevant characteristics of the participants. TBI severity was classified using available clinical and self-report information, including loss of consciousness, post-traumatic amnesia, Glasgow Coma Scale score, and neuroimaging findings. Moderate-to-severe TBI was defined by at least one of the following: post-traumatic amnesia >24 hours, loss of consciousness >30 minutes, Glasgow Coma Scale <13, or structural neuroimaging abnormality. Participants were not excluded solely because one or more severity indicators were unavailable, provided sufficient clinical information supported classification. Missingness in severity indicators was reported descriptively. The primary olfactory outcomes, loss of consciousness, and number of injuries were complete for all TBI participants, and time since last TBI was missing for one participant.

Olfactory function was assessed using standardized odor identification, odor discrimination, and odor memory measures. Odor stimuli were delivered using portable Whispi devices, with odors provided by International Flavors and Fragrances, and study data were recorded in REDCap. Primary analyses used general linear models to test associations between TBI severity and olfactory outcomes, adjusted for age, age², sex, and education. Secondary TBI-only models examined loss of consciousness, number of injuries, and time since last TBI, with additional adjustment for relevant injury-related covariates. Type III tests were used for omnibus effects, and Tukey-adjusted post hoc comparisons were applied when appropriate. Model diagnostics were inspected to evaluate residual distributions and influential observations. Sex as a biological variable was explicitly considered and included as a covariate in all primary models. Limitations related to sample size, sample diversity, injury heterogeneity, and incomplete availability of some clinical severity indicators are described in the Discussion. Analyses were conducted in R using general linear models, Type III tests, estimated marginal means, and Tukey-adjusted comparisons. Data may be requested from the corresponding author, subject to institutional approval and ethical restrictions related to participant confidentiality. Artificial intelligence tools were used to assist with manuscript editing and formatting. The authors reviewed all AI-assisted content and take full responsibility for the accuracy and integrity of the manuscript. The authors acknowledge that inappropriate AI-use will be grounds for rejection or retraction.

## Data Availability

All data produced in the present study are available upon reasonable request to the authors.

## ACKNOWLEDGMENT

We thank the participants in this study. We thank Scentovations and International Flavors and Fragrances for providing Whispis and odors for this study. We thank Sean Reineke and Joseph Lacasio for consultation.

## CRediT AUTHOR CONTRIBUTIONS

A.D.A., R.Z., and M.W.A.: Conceptualization, Methodology.

A.D.A., V.T., A.M., and N.B.: Investigation, Data Curation.

A.D.A. and B.J.: Formal Analysis, Visualization.

A.D.A, B.J., C.R., and D.D.: Writing - Original Draft.

All authors: Writing - Review & Editing.

R.Z. and M.W.A.: Funding Acquisition.

## DATA AVAILABILITY STATEMENT

Data may be requested by contacting the corresponding author M.W.A.

